# Disease Burden and Direct Health-Care Spending on Brain Conditions in Switzerland: Findings from the Global Burden of Disease 2023 Study for the Implementation of the Swiss Brain Health Plan

**DOI:** 10.64898/2026.05.01.26352201

**Authors:** Indrit Bègue, Lorina Sinanaj, Xaviera Steele, Raphael Guzman, Luca Crivelli, Alexandre N. Datta, Claudio L.A. Bassetti

## Abstract

**Background:** Brain disorders are leading contributors to increasing disability and spending worldwide. In 2022 the Swiss Brain Health Plan (SBHP) was launched to promote brain health and prevent brain disorders. To guide the implementation of the SBHP, we performed a detailed analysis of the health and economic burden of brain disorders in Switzerland.

**Methods:** We analyzed Global Burden of Disease 2023 disability-adjusted life years (DALYs) and Institute for Health Metrics and Evaluation (IHME) cause-specific health-care spending estimates for Switzerland. DALYs were quantified for 1990 – 2023. Spending was analyzed for 2000 – 2019 across six types of care. We examined age and sex patterns, spending distribution, and international comparisons with six other countries (Germany, France, Denmark, Norway, Italy, Singapore). To assess short- and longer-term association between burden and spending estimates, we fitted panel regression models with disorder and year fixed effects under one-year and five-year lag specifications.

**Findings:** Both disease burden and spending were highly concentrated in a small number of conditions in Switzerland. In 2023, ten brain disorders accounted for 82·9% of Switzerland’s total DALY burden. In 2019, ten brain disorders accounted for 86·0% of all direct brain-health spending, with dementia alone comprising 29·5% of total expenditures. Among seven analyzed comparator countries, Switzerland had the highest per-capita brain-health spending and the highest spending per DALY. In fixed-effects panel models that accounted for spending persistence, lagged DALYs were not statistically associated with subsequent spending. Suicide prevention and addiction showed significant lower-than-expected health-sector spending (self-harm: β = -0·23; drug use disorders: β = -0·08 to -0·18 across lag models).

**Interpretation:** Brain disorders generate a large burden in Switzerland. Within the IHME estimates, the burden–spending relationship over time appears limited. The implementation of the SBHP will refer to the current data and call for a burden-informed financing to guide strategic cross-sectorial allocation and prevention investments.

**Research in context:** *Evidence before this study:* We drew on evidence from the Global Burden of Disease (GBD) 2023 estimates on neurological and mental health conditions, and on cause-specific health-care spending data from the Institute for Health Metrics and Evaluation for Switzerland and selected high-income countries. These sources show that brain disorders are major contributors to disability and premature mortality, and that Switzerland is among the world’s highest spenders per capita on health care. Prior work has described the costs of individual brain disorders and drivers of health expenditure growth; however, it has often treated burden and spending as partly separate domains, leaving the country-level link between cause-specific disability-adjusted life-years (DALYs) and cause-specific spending, over time and in either direction, poorly characterized.

*Added value of this study:* To our knowledge, this is the first study in a single country to systematically link cause-specific DALYs and cause-specific direct health-care spending for brain disorders and to examine their longitudinal and bi-directional associations. Using harmonized GBD 2023 estimates and IHME 2019 cause-specific health spending data, we quantify the health and economic burden of 23 brain disorders in Switzerland across age, sex, care setting, and time, and benchmark patterns against six other high-income countries. By applying panel regression models with disorder and year fixed effects, we assess whether modeled spending shows any association with prior modeled burden once spending persistence is accounted for, and identify conditions with higher or lower spending relative to burden.

*Implications of all the available evidence:* Switzerland bears a major burden of brain disorders and devotes substantial resources to their care, yet within the modeled estimates, spending does not consistently correspond to burden over time. Disorders with long-standing multisectoral programs tended to show lower spending-to-burden ratios, suggesting that coordinated action beyond the health sector may reduce downstream health-sector demand. For Switzerland and similar health systems, these findings support national brain-health strategies that strengthen life-course prevention and early intervention, and that integrate financing with burden data to inform priority setting and periodic reassessment of resource allocation.

## Introduction

Brain disorders represent a substantial and growing component of the global disease burden, affecting hundreds of millions of people worldwide and profoundly impacting health systems, economies, and society.^1,2^ The World Health Organization and recent Global Burden of Disease studies have documented that neurological and mental health conditions collectively rank among the leading causes of disability and premature mortality across high-income nations.^2,3^ Yet despite clear evidence of their health impact, important questions remain about how health-care spending relates to disease burden over time and how prevention and early intervention might shape future burden and resource needs.^4^

The association between disease burden and health-care spending has important implications for health policy and equity. Ideally, an effective burden-responsive health system would direct a greater share of resources toward conditions associated with the highest levels of disability, provided that such investments generate net reductions in population health burden. On the contrary, when investments are ineffective, increased spending may fail to reduce, and may even worsen, population health outcomes.^5^ However, research in various health-care contexts suggests that spending patterns often reflect historical precedent and entrenched institutional structures rather than current epidemiological evidence^6^. Characterizing how cause-specific burden and cause-specific spending relate over time can therefore inform planning and priority setting. Switzerland represents an instructive case study for examining this association. As a high-income nation with one of the world’s highest per-capita health-care spending, Switzerland provides an opportunity to investigate how a well-resourced system allocates funds for brain disorders. At the same time, Switzerland’s demographic - aging - profile and rising neurological and mental health demand mirror trends emerging in many countries, making its experience informative beyond its borders.

The current study was conceptualized in the context of implementing the Swiss Brain Health Plan (SBHP), launched in 2022 as a holistic, intersectoral national initiative to promote brain health and prevent brain disorders.^7^ The SBHP is motivated by three insights: 1) increasing disability and mortality related to brain disorders, 2) expanding opportunities for prevention, and 3) recognition of brain health as foundational to individual and societal potential. In this context, up-to-date data on the health and economic burden of brain disorders are crucial for implementation and planning.^8^

Building on earlier work using different methodologies^9^, this study has three aims: first, to quantify the burden of brain conditions in Switzerland; second, to characterize direct health-care spending for these conditions; and third, to examine the association between burden and spending over time. We used the most recent Global Burden of Disease data (GBD 2023) together with the latest cause-specific health-care spending estimates (IHME 2019). In addition, we situate Switzerland’s burden and spending patterns in an international context to support interpretation. To our knowledge, this is the first study to longitudinally link annual cause-specific burden for brain disorders with annual cause-specific direct health-care spending in a single country. This analysis aims to inform SBHP implementation, including resource planning for brain disorders in Switzerland, and to provide a framework that other countries can adapt when examining burden and spending together.

## Methods

### Study Design

An overview of the study design is presented in Figure 1A. We conducted a secondary analysis of data from the Global Burden of Disease (GBD) 2023 study^10^ and the Institute for Health Metrics and Evaluation (IHME) health-care spending database.^11^ GBD provides standardized estimates of disability-adjusted life-years (DALYs) by cause, age, sex, location, and year for 1990 – 2023. This analysis leverages newly available direct health-care spending data, complementing GBD burden estimates. IHME reports direct health-care spending by cause, age, sex, location, year, and type of care for 2000 – 2019. Detailed descriptions of both estimation frameworks are available in the original publications.^10, 11^

**Figure 1.**
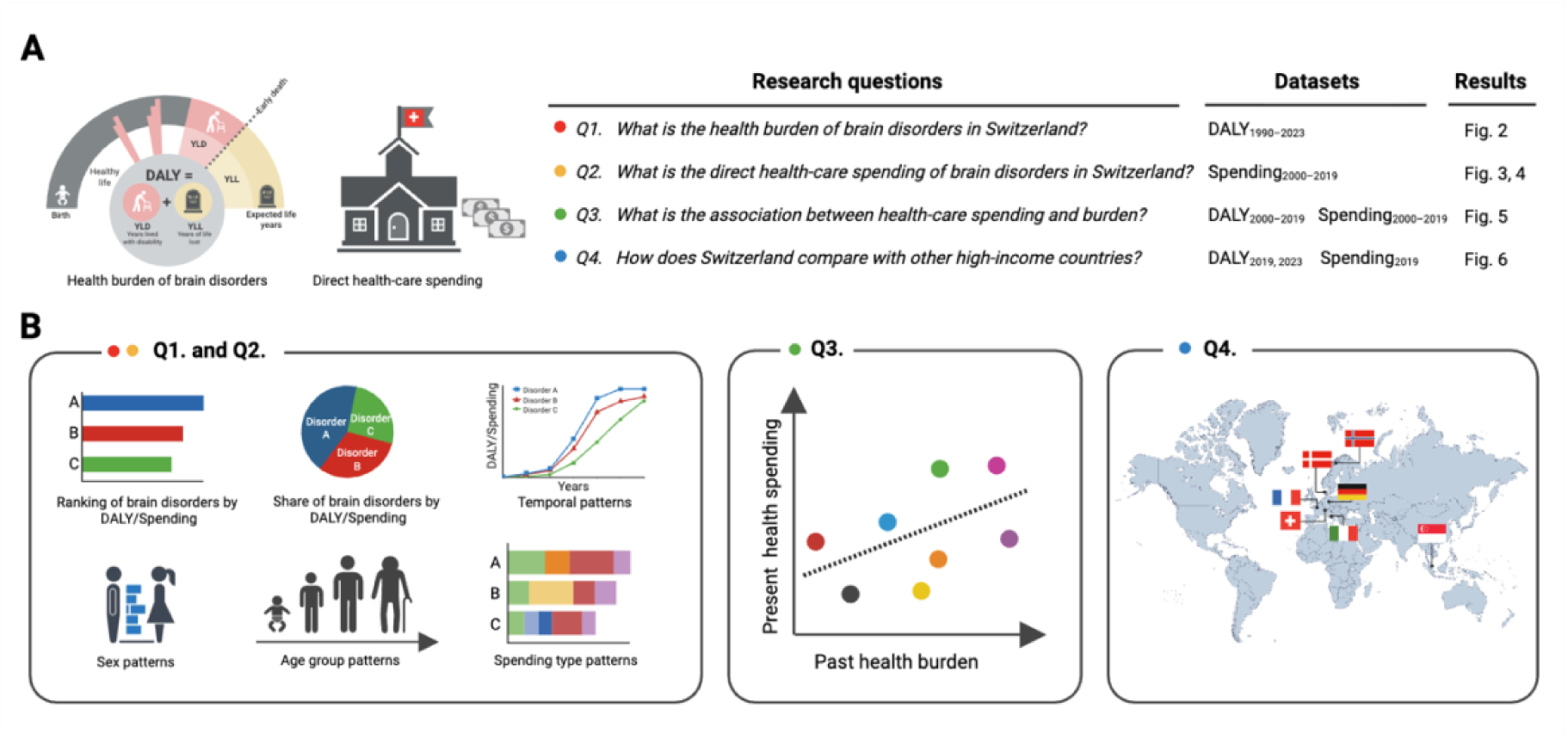
Analytical framework of disease burden and health-care spending in Switzerland. (A) *Study design.* Modeled DALY estimates from the Global Burden of Disease study (1990 – 2023) and modeled direct health-care spending estimates from IHME (2000 – 2019) were integrated to evaluate brain disorders across four analytical domains: health burden, direct health-care spending, temporal association between spending and burden, and international comparison. (B) *Statistical analyses.* Methods applied for each domain included ranking of brain disorders by burden and spending, temporal and demographic analyses, fixed-effects panel regression models assessing the association between spending and burden, and benchmarking Switzerland against France, Germany, Denmark, Norway, Italy, and Singapore. DALY=disability-adjusted life-year.

We examined disease burden and direct health-care spending in Switzerland (Figure 1B, 1C). Brain disorders were defined following IHME’s cause hierarchy^11^ and included 24 conditions: alcohol use disorders; Alzheimer’s disease and other dementias; anxiety disorders; attention-deficit/hyperactivity disorder; autism spectrum disorders; bipolar disorder; brain and central nervous system cancer; conduct disorder; depressive disorders; drug use disorders; eating disorders; encephalitis; headache disorders; idiopathic developmental intellectual disability; idiopathic epilepsy; meningitis; motor neuron disease; multiple sclerosis; other mental disorders; other neurological disorders; Parkinson’s disease; schizophrenia; self-harm; and stroke. Spending estimates were available for 23 of these conditions: opioid and non-opioid drug use disorders were aggregated into a single “drug use disorders” category to ensure comparability with DALY estimates and “other neurological disorders” lacked spending data. We are aware of the fact that a few brain disorders (e.g. sleep disorders, vestibular disorders or traumatic brain disorders, to name a few) were not included in the list of disorders used in the GBD.

DALYs were analyzed for 1990 – 2023. They are defined as the sum of years lived with disability (YLDs) and years of life lost (YLLs) due to premature mortality. Direct health-care spending was analyzed for 2000 – 2019 and harmonized to 2021 US dollars using IHME conversion methods. For Switzerland-specific analyses, we used total DALY count and total spending; for international comparisons, we used aggregated age-standardized DALY rates (per 100,000) to compare burden across countries, and per-capita spending (US$/person; all ages) to compare spending levels. Spending per DALY was calculated for 2019 as per-capita spending divided by the all-ages DALY rate expressed on a per-person basis. Although PPP-adjusted values are often used for cross-country comparisons, we report spending in 2021 US dollars in this secondary analysis as this is the available metric used in the GBD health-spending dataset. To benchmark Switzerland, we selected six comparator countries: Germany, France, Denmark, Norway, Italy and Singapore (Figure 1D). We included Singapore as a non-European country for two main reasons. First, Singapore closely resembles Switzerland in key structural drivers of brain-health burden and spending, including high income, rapid population ageing, and a high-performing, mixed-financing health system. Second, Singapore has translated these challenges into concrete national policies and long-term care strategies. This makes it a relevant implementation benchmark for the Swiss Brain Health Plan and situates Switzerland and its European peers within the broader landscape of high-performing brain-health systems.

### Burden of disease and direct health-care spending analyses

We ranked brain disorders by total DALYs (2023) and total spending (2019) to identify leading contributors. Temporal trends in burden and spending were examined over their respective observation periods. To assess life-course patterns, DALYs and spending were stratified by 5-year age groups (from early neonatal to 85 years and older). Sex-specific analyses examined differences in burden and spending across age groups and conditions. Spending was further disaggregated by six types of care (ambulatory, emergency department, home health, inpatient, nursing facility, and pharmaceuticals). Uncertainty in estimates is reported using 95% uncertainty intervals (UI).

### Association between burden of disease and direct health-care spending

To assess short- and longer-term association between modeled burden and modeled spending, we estimated fixed-effects panel regression models across 23 brain disorders (Figure 1C). We specified two ordinary least squares (OLS) models with condition and year fixed effects: (OLS 1) a one-year lag model (2001 – 2019), in which current spending was modeled as a function of prior-year DALYs and prior-year spending, and (OLS 2) a five-year prior-window average model (2005 – 2019), in which current spending was modeled as a function of five-year averages of prior DALYs and spending. To examine the reverse direction (i.e., whether subsequent DALY estimates are associated with prior spending), we estimated a complementary model with DALYs as the outcome and lagged spending as the predictor, controlling for lagged DALYs and fixed effects (equations in Appendix S1). Because both DALYs and spending are derived from related IHME modeling frameworks, coefficients are interpreted as descriptive elasticities within a modeled system rather than causal. Given any potential for nonnormal distribution of these variables, all were log transformed prior to regression analysis. To account for within-disorder serial correlation and heteroskedasticity, standard errors were clustered at the disorder level. Multicollinearity between regressors (i.e., lagged DALYs and lagged spending) was assessed using pairwise correlations and variance inflation factors (VIF). Throughout this manuscript, analyses with regressors of VIF below 2 are reported, indicating no concern regarding multicollinearity.

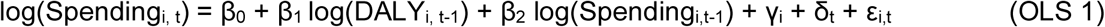

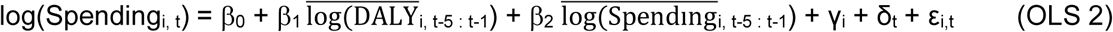

Where: *i* = brain disorder index, *t* = year index*, β_0_* = intercept (baseline log-spending level), *β_1_ =* elasticity of current spending with respect to past DALYs, *β_2_ =* elasticity of current spending with respect to past spending, *γ =* condition fixed effect, *δ =* year fixed effect, *ε =* error term. Model performance was assessed using adjusted R². Statistical significance was evaluated using two-sided tests with α=0·05. All analyses were conducted using Python (version 3·10) with the statsmodels package.

## Results

### Health burden of brain disorders in Switzerland

In 2023, ten brain disorders accounted for 82·9% of the total DALY burden in Switzerland (Figure 2A), led by Alzheimer’s disease and other dementias (100 691·4 DALYs [95% UI 44 301 – 212 704]; 15·4% of the total burden), depressive disorders (94 630·4 DALYs [62 905 – 135 492]; 14·4%) and anxiety disorders (84 878·9 DALYs [47 268 – 146 836]; 12·9%). Full numerical estimates with uncertainty intervals for all 24 disorders are provided in Table S1.

**Figure 2.**
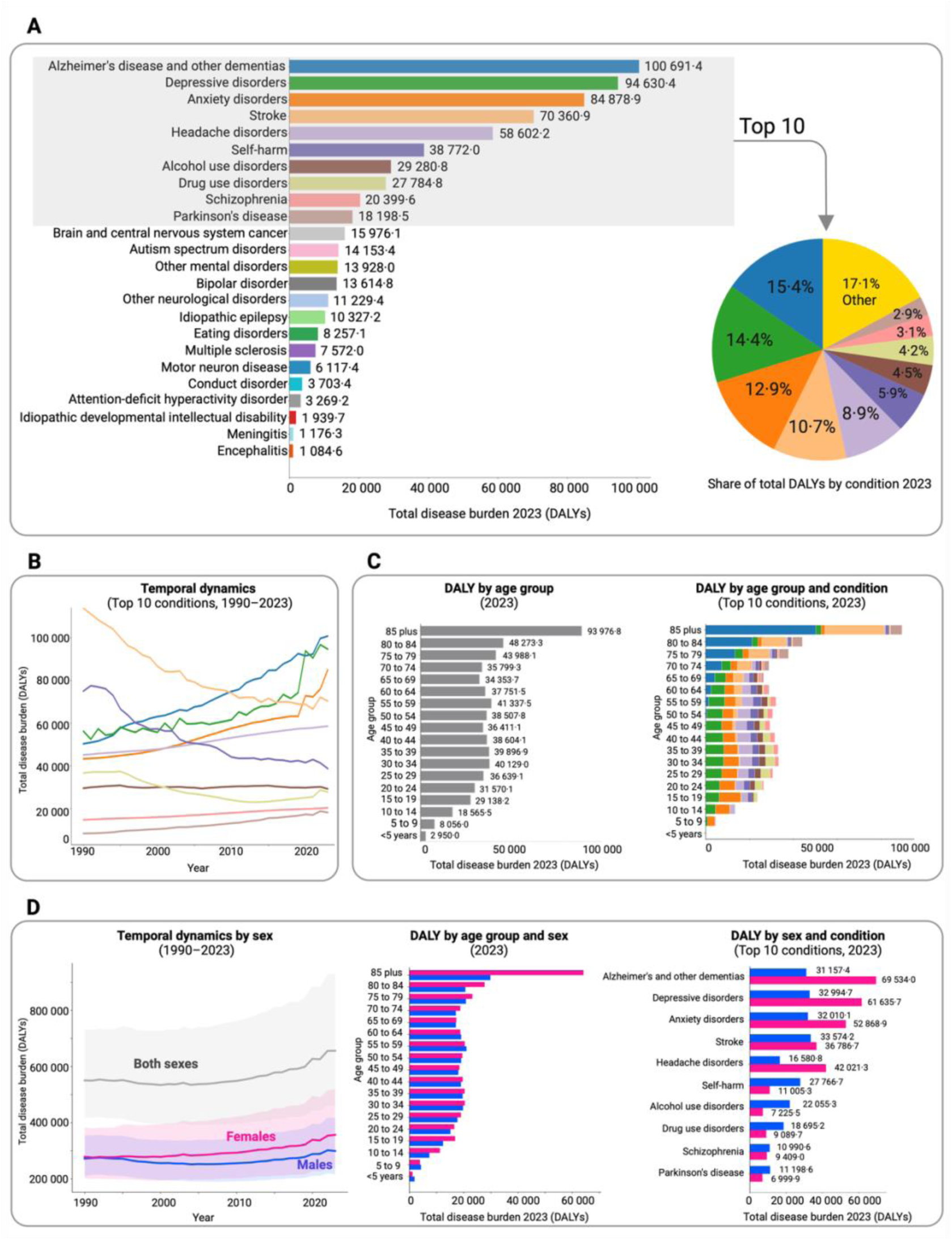
Health burden of brain disorders in Switzerland, 1990 – 2023. (A) Ranking of brain disorders by total DALYs in 2023 and proportion of total burden contributed by the ten leading causes. (B) Temporal trends in total DALYs for the top ten disorders, 1990 – 2023. (C) Distribution of DALYs by age group and condition in 2023. (D) DALY trends by sex (1990 – 2023) and by age group and condition in 2023. DALYs=Disability-Adjusted Life-Years.

From 1990 to 2023, dementia showed the largest increase in DALY burden, whereas stroke, self-harm, and drug use disorders declined (Figure 2B). Projections to 2050 based on forecasted GBD 2021 estimates (most recent GBD with available projections to 2050)^2^, indicate further growth in dementia burden, with most other conditions remaining stable or showing only modest increases (Figure S1).

Age-stratified analyses showed that in 2023, DALY burden increased progressively with age, peaking among individuals aged 85 years and older. Mental disorders (particularly depressive and anxiety disorders) dominated burden in early and mid-adulthood, while more neurological disorders, especially dementia and stroke, dominated in older age groups (Figure 2C).

Sex-stratified analyses showed that women consistently carried a higher DALY burden than men across 1990 to 2023 (Figure 2D). In 2023, total DALYs were 356 660 [268 016 – 445 035] in women and 299 288 [251 325 – 352 224] in men. The largest differences were observed during biological transitions (puberty, peripartum, perimenopause and menopause period) and after age 75 (Figure 2D). Headache disorders were 2·53 times higher in females, followed by eating disorders (2·34 times higher) and dementia (2·23 times higher), whereas male burden was higher for autism spectrum disorders (3·25 times higher in males), alcohol use disorders (3·05 times higher) and self-harm (2·52 times higher). Full sex differences in DALYs across brain disorders are shown in Figure S2.

### Economic burden of brain disorders in Switzerland

In 2019, ten brain disorders accounted for 86% of the total direct health-care spending in Switzerland (Figure 3A), led by Alzheimer’s disease and other dementias (total mean spending $5 969·8 million [2 540·9 – 15 193·2]; 29·5% of total brain disorder spending), anxiety disorders ($2 204·9 million [1 910·4 – 2 599·4]; 10·9%) and stroke ($2 169·1 million [1 428·4 – 3 351·8]; 10·7%). Mean and uncertainty intervals for all 23 disorders are shown in Table S2.

**Figure 3.**
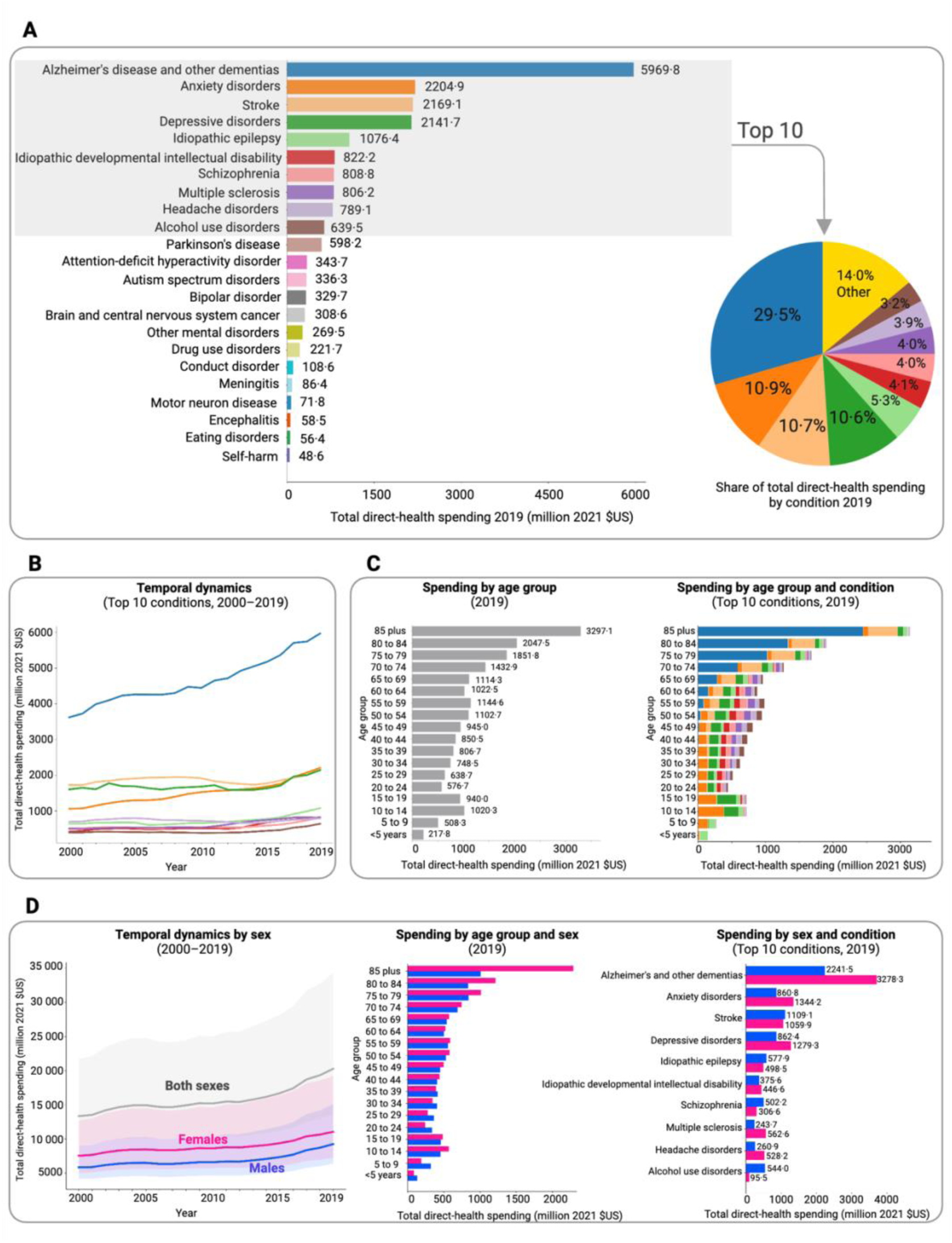
Direct health-care spending on brain disorders in Switzerland, 2000 – 2019. (A) Ranking of brain disorders by total direct spending in 2019 and proportion of total spending contributed by the ten leading causes. (B) Temporal trends in total spending for the top ten disorders, 2000 – 2019. (C) Distribution of spending by age group and condition in 2019. (D) Spending trends by sex (2000 – 2019) and by age group and condition in 2019.

Across 2000 – 2019, total spending increased steadily, with Alzheimer’s disease and other dementias showing the largest rise (Figure 3B). Age-stratified results showed the highest spending among individuals aged 85 years and older. Spending in younger age groups (aged 10 – 19 years), driven largely by depressive and anxiety disorders, was comparable to that of middle-aged adults (aged 60 – 64 years), where dementia and stroke accounted for most costs (Figure 3C).

Sex-stratified analyses showed that total direct care spending was higher for women than men from 2000 to 2019 (Figure 3D). In 2019, total spending for brain disorders in Switzerland reached $20·3 billion [15·5 – 31·6], with females accounting for $11·0 billion [8·3 – 17·9] and males for $9·2 billion [7·2 – 13·7]. Female spending exceeded male spending in most age groups, with particularly high differences after age 75. The largest female-to-male ratios were observed for eating disorders (4·70 times higher in females), multiple sclerosis (2·31 times higher), and headache disorders (2·02 times higher), whereas men had higher spending for alcohol use disorders (5·70 times higher), autism spectrum disorders (4·06 times higher), and drug use disorders (2·17 times higher). Full sex differences in spending across brain disorders are shown in Figure S3.

Spending type varied markedly by disorder (Figure 4). The largest share of total spending was directed to nursing facility care, accounting for 85% of spending in Alzheimer’s disease and other dementias. Inpatient care was the primary spending driver for stroke (52%), depressive disorders (40%), schizophrenia (50%), headache (37%) and alcohol use disorders (68%). Ambulatory care accounted for the largest share of spending for anxiety disorders (45%), while prescribed pharmaceuticals were the primary driver for multiple sclerosis (45%). Female health-care spending exceeded that of males across ambulatory services and for pharmaceutical services, whereas inpatient spending was higher among males (Figure S4).

**Figure 4.**
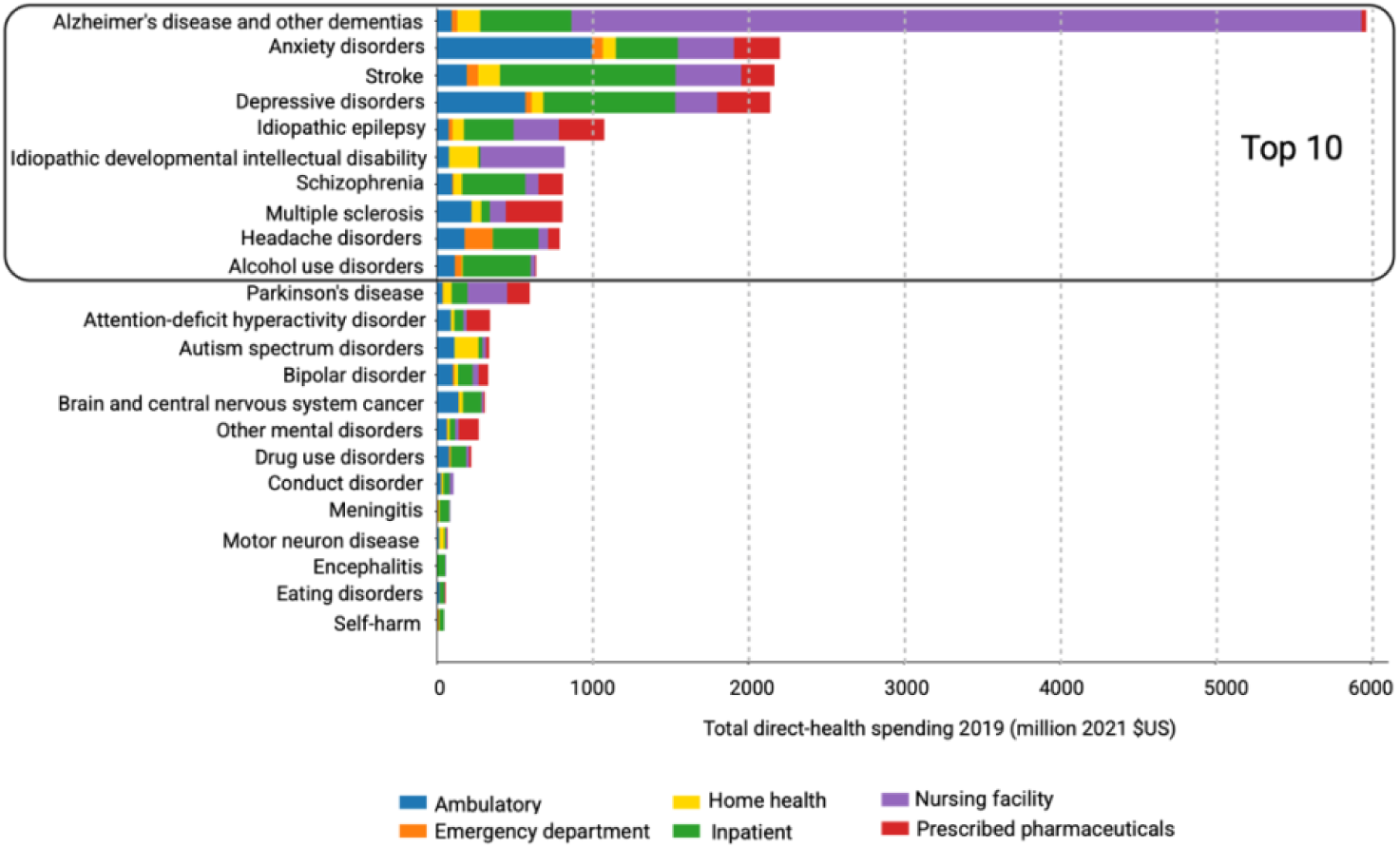
Distribution of direct health-care spending across care types for brain disorders in Switzerland in 2019. Stacked bars show condition-specific composition of total spending across six care types: ambulatory, emergency department, home health, inpatient, nursing facility, and pharmaceuticals.

### Switzerland shows limited temporal association between burden and spending

Fixed-effects panel regressions showed that, after accounting for spending persistence and disorder and year fixed effects, lagged DALYs were small in magnitude and not statistically associated with subsequent spending in either the one-year or five-year specification (Table 1; Figure 5A, 5B). In complementary reverse-regression analyses, DALYs were highly persistent, and lagged spending did not show any association with year-ahead DALY estimates (Table S3).

**Table 1:** Panel estimates of the association between disease burden and health-care spending for 23 brain disorders in Switzerland, 2001-2019.

|  | Model 1 | Model 2 |
| --- | --- | --- |
| Years window | 1 | 5 |
| log(DALY) | 0·078 (0·062) | 0·17 (0·19) |
| log(Spending) | 0·93 (0·033)** | 0·88 (0·12)** |
| Observations (conditions x year) | 437 (23 x 19) | 345 (23 x 15) |
| R<sup>2</sup> | 0·9990 | 0·9975 |
| Adjusted R<sup>2</sup> | 0·9989 | 0·9971 |
| Abbreviations: DALY, disability-adjusted life-years.<br>Standard errors clustered by condition. Coefficients should be interpreted as elasticities.<br>Constants are excluded. ** p<0·001. |  |  |

**Figure 5.**
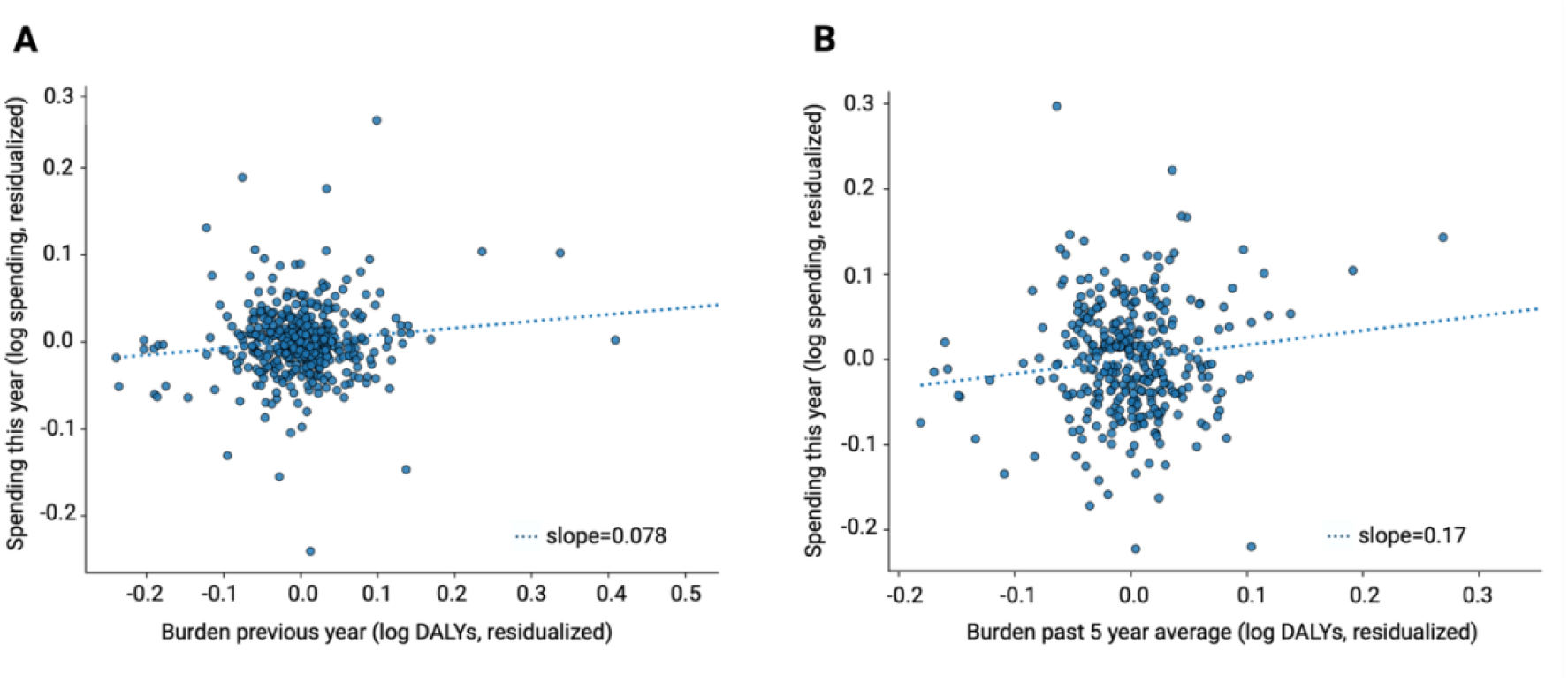
Association between health-care spending and prior disease burden in Switzerland. (A) *One-year lag model (2001 – 2019).* Fixed-effects panel regression estimating the association between current spending and prior-year DALYs, adjusting for prior-year spending and disorder and year fixed effects. (B) *Five-year window average* model (2005 – 2019). Fixed-effects panel regression estimating the association between current spending and five-year averages of prior DALYs, adjusting spending persistence and disorder and year fixed effects. DALYs=Disability-Adjusted Life-Years.

### Lower spending relative to burden for self-harm and drug use disorders in Switzerland

Condition-level fixed effects captured systematic deviations in disorder-specific spending not captured by disease burden or historical spending. Detailed coefficients for fixed effects are presented in Table S4 (one-year lag model) and Table S5 (five-year lag model). Drug use disorders showed consistently negative and statistically significant fixed effects in both models. In the one-year lag model, drug use disorders deviated negatively from expected spending (β = −0·078, clustered SE = 0·014, p < 0·001), and this negative deviation remained significant in the five-year lag model (β = −0·18, clustered SE = 0·059, p = 0·002). In the one-year lag model, self-harm also showed a significant negative deviation from expected spending (β = −0·23, clustered SE = 0·089, p = 0·008), while Alzheimer’s disease and other dementias and anxiety disorders demonstrated higher modeled spending (β = 0·11, clustered SE = 0·051, p = 0·036, and β = 0·06, clustered SE = 0·027, p = 0·036, respectively). Overall, these fixed effects indicate that modeled spending remains systematically lower for self-harm and drug use disorders and modestly higher for Alzheimer’s disease and other dementias and anxiety disorders. Year fixed effects showed a structural shift in national brain-health spending after 2015 (Appendix S2).

### Switzerland has the highest spending per unit of DALY burden in cross-country comparisons

Switzerland’s aggregated age-standardized DALY rate for brain disorders in 2023 (5708·9 per 100 000 population, 95% UI 4608·6 – 6936·1) was lower than Germany, France, Norway and Denmark, but higher than Italy and Singapore (Figure 6A). In 2019, Switzerland had the highest aggregated per-capita brain-health spending (US$ 2331·2, 95% UI 1779·5 – 3634·5) among the compared countries (Figure 6B). When combining 2019 per-capita spending with 2019 DALY rates, Switzerland also had the highest spending per unit of DALY burden (US$ 33 837 per DALY), compared with US$ 31 269 in Norway, US$ 19 570 in Denmark, US$ 18 876 in Germany, US$ 14 985 in France, US$ 8628 in Italy, and US$ 6553 in Singapore (Figure 6C). A detailed breakdown by condition (Figure S5) suggests this pattern is largely driven by high spending on Alzheimer’s disease, stroke, and depressive and anxiety disorders.

**Figure 6.**
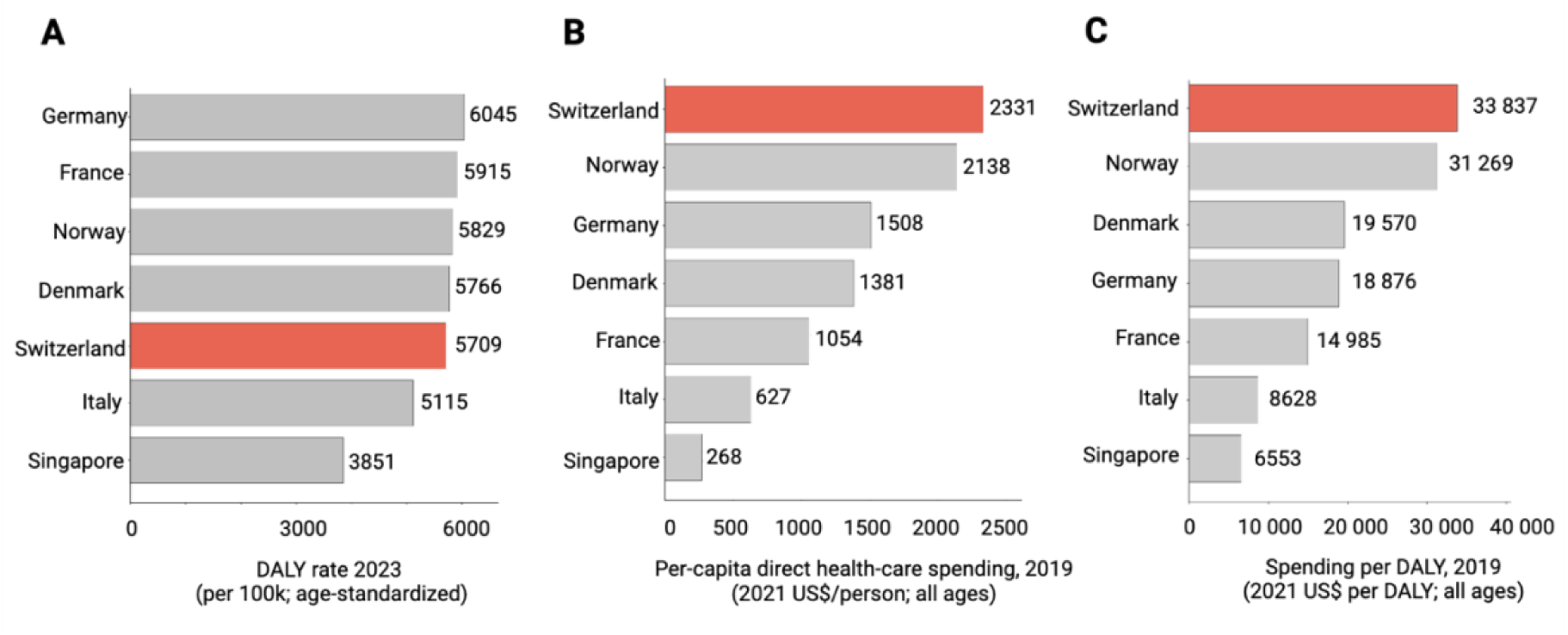
International comparison of burden and spending across high-income countries. (A) Age-standardized DALY rates for brain disorders in 2023. (B) Per-capita direct health-care spending for brain disorders in 2019 (2021 US$/person; all ages). (C) Spending per DALY in 2019, calculated as per-capita spending 2019 divided by the all-ages DALY rate 2019 converted to a per-person rate (2021 US$/DALY; all ages). DALYs=Disability-Adjusted Life-Years. IHME=Institute for Health Metrics and Evaluation. GBD=Global Burden of Disease.

## Discussion

This study shows that brain disorders account for a substantial share (24%) of health loss and direct health-care expenditure in Switzerland. The burden is large and unevenly distributed across the life course and between sexes, reflecting both biological vulnerability and social determinants.^12^ Mental disorders predominate in adolescence, young adulthood, and midlife – periods marked by neurodevelopmental transitions and heightened sensitivity to environmental stressors^13^ – whereas most neurological disorders, particularly dementia and stroke, appear to dominate in older age as neurodegenerative and cerebrovascular risks accumulate.^14^ These life-course patterns are consistent with earlier European cost-of-illness estimates^9^ and with recent national registry-based analyses^15^, similarly documenting a lifespan-specific concentration of disability and costs, although direct quantitative comparison is limited by differences in disease groupings, costing methods, and outcome definitions. Sex differences were striking across mental and neurological disorder domains (Figures 2D, 3D, S1, S2) and reflect the importance of sex-sensitive prevention and access to care planning when interpreting burden – spending patterns. Our analyses align well with a recent Swiss National Health Accounts decomposition.^21^ In 2017, Stucki and colleagues^21^ attributed 11·37 billion CHF to mental disorders and 6·75 billion CHF to neurological disorders (18·12 billion CHF combined), which is comparable in magnitude to our 2019 estimates of $20·3 billion (2021 USD). Despite differences in year, currency, and attribution methods, care-type patterns are also consistent across approaches: to illustrate, both reports allocate the majority of dementia spending to long-term/nursing facility care (82 – 85%), show inpatient care as the dominant component in similar conditions (e.g., schizophrenia, substance use disorders), and highlight pharmaceuticals as a large share of the economic burden in similar context (e.g., multiple sclerosis). Together, these findings underscore that brain disorders (their prevention as well as promotion of brain health) is a major public-health priority for Switzerland and advocate for a coordinated response that aligns with life-course, sex-informed biology and long-term trajectory of these conditions.^7,16^

Across both lag specifications, we found limited evidence that changes in disease burden are associated with subsequent changes in health-care spending after accounting for spending persistence and fixed effects (Figure 5). In addition, lagged spending was not associated with subsequent burden. Although these analyses bear no significance on causality, this pattern reinforces the concern that higher expenditure does not necessarily translate into population health gains but may, in contrast, have the opposite effect by displacing more effective uses of resources^5^. To illustrate, increased National Health Service (NHS) spending in England on high-cost cancer drugs displaced more population health than it produced during 2000 – 2020.^5^ Had these resources instead been invested in existing NHS services, an estimated 5·0 million additional quality-adjusted life-years (QALYs) could have been generated. An additional point of reflection stems from the observation of spending inertia, which may be reflective of structural features of the Swiss health-care system. Responsibilities for planning, financing, and regulation are split across federal authorities, cantons, and more than 50 insurers.^17^ Such configuration creates strong path dependence and limits rapid reallocation in the face of epidemiological changes. Taken together, these findings suggest that burden-informed allocation needs to be coupled with evidence on effectiveness and value, rather than merely assuming that increased spending alone will reduce disability. In addition, they also strengthen the case for prevention and early intervention as key levers to reduce future demand. In fact, long-term analyses of Swiss health-care expenditure over the period 1960 – 2022 suggest that system-wide drivers, such as population ageing and workforce constraints amongst many others, apply sustained upward pressure on spending.^18^ Therefore, investment in prevention is not only a public-health priority but also one of the few realistic tools to ease future pressure on a system that has limited flexibility to reallocate resources. In keeping with this, the lower-than-expected health-sector spending observed for self-harm and drug use disorders (Table S4, S5) could reflect the beneficial effects of long-standing national programs in suicide prevention and addiction in Switzerland geared toward prevention, early detection, harm reduction and education. Because much of this activity typically lies outside the formal health-care budget, the budget expenditure estimates here focused on *medical* spending are positively associated with burden: lower direct health-sector spending in these conditions likely reflects the success of upstream prevention and coordinated action and lowered burden. These conditions highlight the potential of multisector prevention for other brain-health domains where fragmentation remains a barrier.

International comparisons reinforce the need for a more coherent approach. Switzerland’s burden of brain disorders is similar to that of neighboring countries,^2,19^ yet its expenditure is substantially higher.^20,21^ This pattern is consistent with Switzerland’s high total health expenditure per capita, which also ranks above Norway, Germany, Denmark, France, Italy and Singapore, suggesting that differences in brain-health spending may partly reflect broader system-wide spending levels.^22^ High wages and comprehensive insurance coverage contribute to this difference,^21^ but the gap also highlights opportunities to improve efficiency, especially in prevention, early intervention, and transitions between levels of care.^23^ Singapore’s Healthier SG health-care delivery strategy reflects this shift from curative care toward preventive, integrated, and community-based care.^24^ Countries that have introduced coordinated brain-health or mental-health strategies – linking governance, financing, and care delivery – have demonstrated improvements in quality, equity, and cost control.^24–26^

Together, these findings support the Swiss Brain Health Plan, an interdisciplinary action plan focused on both neurological and psychiatric disorders by raising awareness, prevention starting in early childhood and encompassing all age groups while supporting patients and caregivers, strengthening interdisciplinary education and research, through the implementation of a holistic and person-centered public-health approach across sectors.^27^ Rather than replacing existing national strategies, the plan provides a shared framework with explicit financial and organizational mechanisms to align them. The relative efficiency observed in suicide prevention^28^ and addiction^29^ illustrates the value of cross-sectoral coordination, where schools, social services, and public-safety systems act alongside health-care to reduce downstream utilization. Finally, a national plan would benefit from integrated data infrastructure linking health-care, long-term-care, social-service, and education datasets, enabling more accurate monitoring, forecasting, and system planning.

This study has strengths and limitations. Strengths include the use of harmonized burden and spending estimates, enabling longitudinal analyses and international benchmarking, and a panel approach that captures movement rather than cross-sectional snapshots. Disorder-level fixed effects help to identify conditions where spending systematically diverges from burden. Because burden and spending data are IHME modeled outputs, and spending estimation incorporates epidemiologic inputs and temporal smoothing, the two series are not statistically independent; regression coefficients should therefore be interpreted as descriptive elasticities within a modeled system. The burden–spending relationship may also be lagged or non-monotonic, so short-run associations may not capture longer-term effects of prevention or chronic disease management.

Other limitations include the exclusion of non-medical and indirect costs (e.g., productivity losses and caregiver burden) and of investments outside the health sector. DALYs may understate severity and social impact for some psychiatric conditions and may not fully capture long-term functional sequelae or comorbidity for neurological disorders. Sleep disorders were not included in this study, even though they affect more than 20% of the general population and were shown on different occasions to be one of the top drivers of costs of neurological disorders^30^ and brain disorders in Europe.^9,15^ Finally, DALYs rely on global disability weights that may not fully reflect the lived experience or societal valuation of disability in Switzerland.

In summary, Switzerland faces a substantial burden of brain disorders that generates high economic costs. Spending patterns remain strongly shaped by historical trajectories rather than current needs, yet areas with established national programs show efficiency. These findings support the implementation of the Swiss Brain Health Plan to align financing, prevention, and care across the life course and across sectors, ensuring that investment reflects the true scale and complexity of brain-health needs.

## Contributors

IB and CB conceptualized this study. IB and XS gained data access and verified the data. LS performed the statistical analyses and created the figures. IB, LS and CB interpreted the results and wrote the manuscript. LS prepared the supplementary material. All authors contributed to writing (review and editing). IB gained funding for the study. IB and CB provided guidance throughout this project. All authors approved the final manuscript and accepted the responsibility for the decision to submit it for publication.

## Data sharing

The data used in these analyses are available online. Global Burden of Disease 2023 estimates can be accessed through the Global Health Data Exchange. Cause-specific health spending estimates from the Institute for Health Metrics and Evaluation are openly available through the IHME health-spending database.

## Acknowledgments

We acknowledge the board and the extended board of the Swiss Brain Health Plan (SBHP), in particular Alice Accorroni and Emiliano Albanese for fruitful discussions in various stages of this work. We acknowledge the Swiss Federation of Clinical Neurosocieties (SFCNS) for the support of the SBHP. The Swiss Brain Health Foundation supports the implementation of the SBHP in Switzerland.

## References

1. Li Y, Jönsson L. The health and economic burden of brain disorders: Consequences for investment in diagnosis, treatment, prevention and R&D. Cereb Circ - Cogn Behav. 2025 Jan 1;8:100377.

2. Ferrari AJ, Santomauro DF, Aali A, Abate YH, Abbafati C, Abbastabar H, et al. Global incidence, prevalence, years lived with disability (YLDs), disability-adjusted life-years (DALYs), and healthy life expectancy (HALE) for 371 diseases and injuries in 204 countries and territories and 811 subnational locations, 1990–2021: a systematic analysis for the Global Burden of Disease Study 2021. The Lancet. 2024 May 18;403(10440):2133–61.

3. Global, regional, and national burden of 12 mental disorders in 204 countries and territories, 1990–2019: a systematic analysis for the Global Burden of Disease Study 2019. Lancet Psychiatry. 2022 Feb 1;9(2):137–50.

4. Vigo DV, Kestel D, Pendakur K, Thornicroft G, Atun R. Disease burden and government spending on mental, neurological, and substance use disorders, and self-harm: cross-sectional, ecological study of health system response in the Americas. Lancet Public Health. 2019 Feb 1;4(2):e89–96.

5. Naci H, Murphy P, Woods B, Lomas J, Wei J, Papanicolas I. Population-health impact of new drugs recommended by the National Institute for Health and Care Excellence in England during 2000–20: a retrospective analysis. The Lancet. 2025 Jan 4;405(10472):50–60

6. Gao T, Han O, Li G, Zeng W. Allocation of the National Institutes of Health funding and burden of disease in 2008–2021 in the United States. Npj Health Syst. 2025 Nov 4;2(1):41.

7. Bassetti CLA, Heldner MR, Adorjan K, Albanese E, Allali G, Arnold M, et al. The Swiss Brain Health Plan 2023–2033. Clin Transl Neurosci [Internet]. 2023 Nov 13 [cited 2026 Jan 9];7(4). Available from: https://www.mdpi.com/2514-183X/7/4/38

8. Bassetti CLA. The Swiss Brain Health Plan in the International Context. Clin Transl Neurosci [Internet]. 2025 Sep 25 [cited 2026 Jan 9];9(4). Available from: https://www.mdpi.com/2514-183X/9/4/44

9. Gustavsson A, Svensson M, Jacobi F, Allgulander C, Alonso J, Beghi E, et al. Cost of disorders of the brain in Europe 2010. Eur Neuropsychopharmacol. 2011 Oct 1;21(10):718–79.

10. Hay SI, Ong KL, Santomauro DF, A B, Aalipour MA, Aalruz H, et al. Burden of 375 diseases and injuries, risk-attributable burden of 88 risk factors, and healthy life expectancy in 204 countries and territories, including 660 subnational locations, 1990–2023: a systematic analysis for the Global Burden of Disease Study 2023. The Lancet. 2025 Oct 18;406(10513):1873–922.

11. Mitchell AJ, Cogswell IE, Dalos J, Tsakalos G, Lei J, Oros A, et al. Estimating global direct health-care spending on neurological and mental health between 2000 and 2019: a modelling study. Lancet Public Health. 2025 May 1;10(5):e401–11.

12. Schrempft S, Trofimova O, Künzi M, Ramponi C, Lutti A, Kherif F, et al. The Neurobiology of Life Course Socioeconomic Conditions and Associated Cognitive Performance in Middle to Late Adulthood. J Neurosci [Internet]. 2024 Apr 24 [cited 2025 Nov 17];44(17).

13. Kretzer S, Lawrence AJ, Pollard R, Ma X, Chen PJ, Amasi-Hartoonian N, et al. The Dynamic Interplay Between Puberty and Structural Brain Development as a Predictor of Mental Health Difficulties in Adolescence: A Systematic Review. Biol Psychiatry. 2024 Oct 1;96(7):585–603.

14. Feigin VL, Vos T, Nichols E, Owolabi MO, Carroll WM, Dichgans M, et al. The global burden of neurological disorders: translating evidence into policy. Lancet Neurol. 2020 Mar 1;19(3):255–65.

15. Vestergaard SV, Rasmussen TB, Stallknecht S, Olsen J, Skipper N, Sørensen HT, et al. Occurrence, mortality and cost of brain disorders in Denmark: a population-based cohort study. BMJ Open. 2020 Nov 1;10(11):e037564.

16. Bègue I, Flahault A, Bolon I, Ruiz de Castañeda R, Vicedo-Cabrera AM, Bassetti CLA. One brain, one mind, one health, one planet—a call from Switzerland for a systemic approach in brain health research, policy and practice. Lancet Reg Health - Eur. 2025 Mar 1;50:101229.

17. Crivelli L, Salari P. The inequity of the Swiss health care system financing from a federal state perspective. Int J Equity Health. 2014;13:17. doi:10.1186/1475-9276-13-17. Available at: 10.1186/1475-9276-13-17

18. Lerch B, Colombier C, Brändle T. Determinants of Health-care Expenditure: Evidence from Switzerland between 1960–2022.

19. Wang Z, Dou Y, Yang X, Guo X, Ma X, Zhou B, et al. Global, regional, and national burden of mental disorders among adolescents and young adults, 1990–2021: a systematic analysis for the Global Burden of Disease Study 2021. Transl Psychiatry. 2025 Oct 10;15(1):397.

20. OECD. Health at a Glance 2025: OECD Indicators – Switzerland. OECD Publishing; 2025. Available from: https://www.oecd.org/en/publications/health-at-a-glance-2025_15a55280-en/switzerland_7139bf0d-en.html

21. Stucki M, Schärer X, Trottmann M, Scholz-Odermatt S, Wieser S. What drives health care spending in Switzerland? Findings from a decomposition by disease, health service, sex, and age. BMC Health Serv Res. 2023 Oct 25;23(1):1149.

22. Global Health Expenditure database, World Health Organization (WHO), uri: apps.who.int/nha/database, publisher: World Health Organization

23. Hughes G, Shaw SE, Greenhalgh T. Rethinking Integrated Care: A Systematic Hermeneutic Review of the Literature on Integrated Care Strategies and Concepts. Milbank Q. 2020;98(2):446–92.

24. The Lancet Regional Health – Western Pacific. Healthier SG: for a healthier Singapore and beyond. Lancet Reg Health West Pac. 2023 Aug 31;37:100893. doi:10.1016/j.lanwpc.2023.100893 PubMed PMID: 37693866; PubMed Central PMCID: PMC10485661.

25. Bassilios B, Currier D, Krysinska K, Dunt D, Machlin A, Newton D, et al. Government funded suicide prevention in Australia – an environmental scan. BMC Public Health. 2024 Aug 26;24(1):2315.

26. Clark DM, Layard R, Smithies R, Richards DA, Suckling R, Wright B. Improving access to psychological therapy: Initial evaluation of two UK demonstration sites. Behav Res Ther. 2009 Nov;47(11):910–20.

27. Bassetti CLA, Bègue I, Chen C, Manes F, Leonardi M, Njamnshi AK, et al. The Bern Declaration on Brain Health: a decalogue to launch an international alliance. Lancet Neurol. 2025 Sep 1;24(9):720.

28. Mack A, Rajkumar S, Kofler J, Wyss K. Estimating the burden of disease attributable to non-assisted suicide in Switzerland from 2009 to 2021: a secondary data analysis. Swiss Med Wkly. 2024 Nov 25;154:3522.

29. Marzel A, Kusejko K, Weber R, Bruggmann P, Rauch A, Roth JA, et al. The Cumulative Impact of Harm Reduction on the Swiss HIV Epidemic: Cohort Study, Mathematical Model, and Phylogenetic Analysis. Open Forum Infect Dis. 2018 May 1;5(5):ofy078.

30. Bassetti CLA, Welter LS, Montes-Martinez M, Mühlberger N, Boon P, Berger T, et al. Epidemiology and economic burden of sleep disorders in Europe. European Journal of Neurology. In press 2026.

