## Supplementary Material for "Disease Burden and Direct Health-Care Spending on Brain Conditions in Switzerland: Findings from the Global Burden of Disease 2023 Study for the Implementation of the Swiss Brain Health Plan"

**Appendix S1:** Reverse-regression analysis models

**Appendix S2:** System-wide health-care spending in Switzerland

**Figure S1:** Top 10 conditions temporal dynamics, Switzerland 1990 – Projected 2050

**Figure S2:** Sex ratio of DALYs by brain disorder, Switzerland 2023

**Figure S3:** Sex ratio of total direct health-care spending by brain disorder, Switzerland 2019

**Figure S4:** Type of spending by age and sex, Switzerland 2019

**Figure S5:** Per-capita spending per DALY by condition and country, 2019

**Table S1:** Health burden mean and uncertainty intervals for 24 brain disorders, Switzerland 2023

**Table S2:** Economic burden mean and uncertainty intervals for 23 brain disorders, Switzerland 2019

**Table S3:** Reverse-regression results

**Table S4:** Coefficients for disorder and year fixed effects in the one-year lag model

**Table S5:** Coefficients for disorder and year fixed effects in the five-year lag model

**Appendix S1: Reverse-regression analysis models**

To assess potential bi-directionality between burden and spending, we estimated a reverse equation in which disease burden (DALYs) is the dependent variable and prior spending is the key predictor. This analysis is intended as a descriptive complement to the main spending-responsiveness models and does not establish causal effects of spending on subsequent burden. OLS 1 is a 1-year lag model (2001-2019) and OLS 2 is a 5-year lag model (2005-2019).

log(DALY_i, t_) = β_0_ + β_1_ log(Spending_i,t-1_) + β_2_ log(DALY_i, t-1_) + γ_i_ + δ_t_ + ε_i,t_	(OLS 1)

log(DALY_i, t_) = β_0_ + β_1_ $\bar{log(Spending}$_i, t-5 : t-1_) + β_2_ $\bar{log(DALY}$_i, t-5 : t-1_) + γ_i_ + δ_t_ + ε_i,t_	(OLS 2)

**Appendix S2: System-wide health-care spending in Switzerland**

Year fixed effects capture system-wide shifts in modeled health-care spending across brain disorders after accounting for disorder baselines, prior burden, and prior spending (with 2001 as the reference year in the one-year model and 2005 in the five-year prior-window model). Detailed coefficients are reported in Supplementary Table S4 (one-year lag model) and Supplementary Table S5 (five-year lag model). In the one-year lag model, spending was lower than expected in 2006 (β = −0.0417, p < 0.001), whereas later years showed consistently higher spending than expected, particularly 2015–2019 (β = 0.0389–0.0600; all p ≤ 0.001). In addition, early positive deviations were observed in 2002–2003 (β = 0.0400 and 0.0248; p < 0.01). The five-year prior-window model showed a similar pattern: significantly negative deviations in 2006–2011 (β = −0.0551 to −0.0305; p ≤ 0.05, with 2011 borderline at p = 0.050), followed by a marked reversal from 2016 onward, with 2016–2019 indicating above-expected spending (β = 0.0698–0.1128; p ≤ 0.015). Together, these year effects are consistent with an upward shift in modeled spending beginning in the mid-2010s, net of disorder-specific changes.

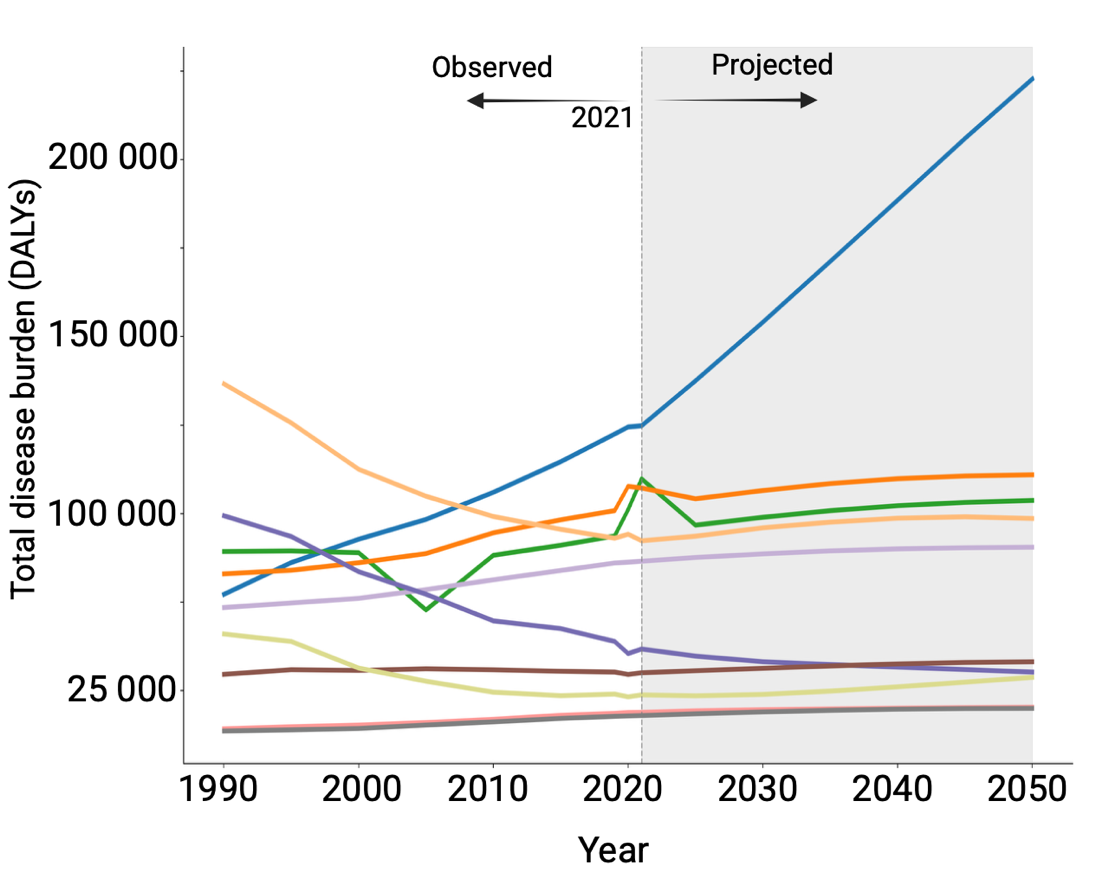

**Figure S1: Top 10 conditions, Temporal dynamics, Switzerland 1990 – Projected 2050**

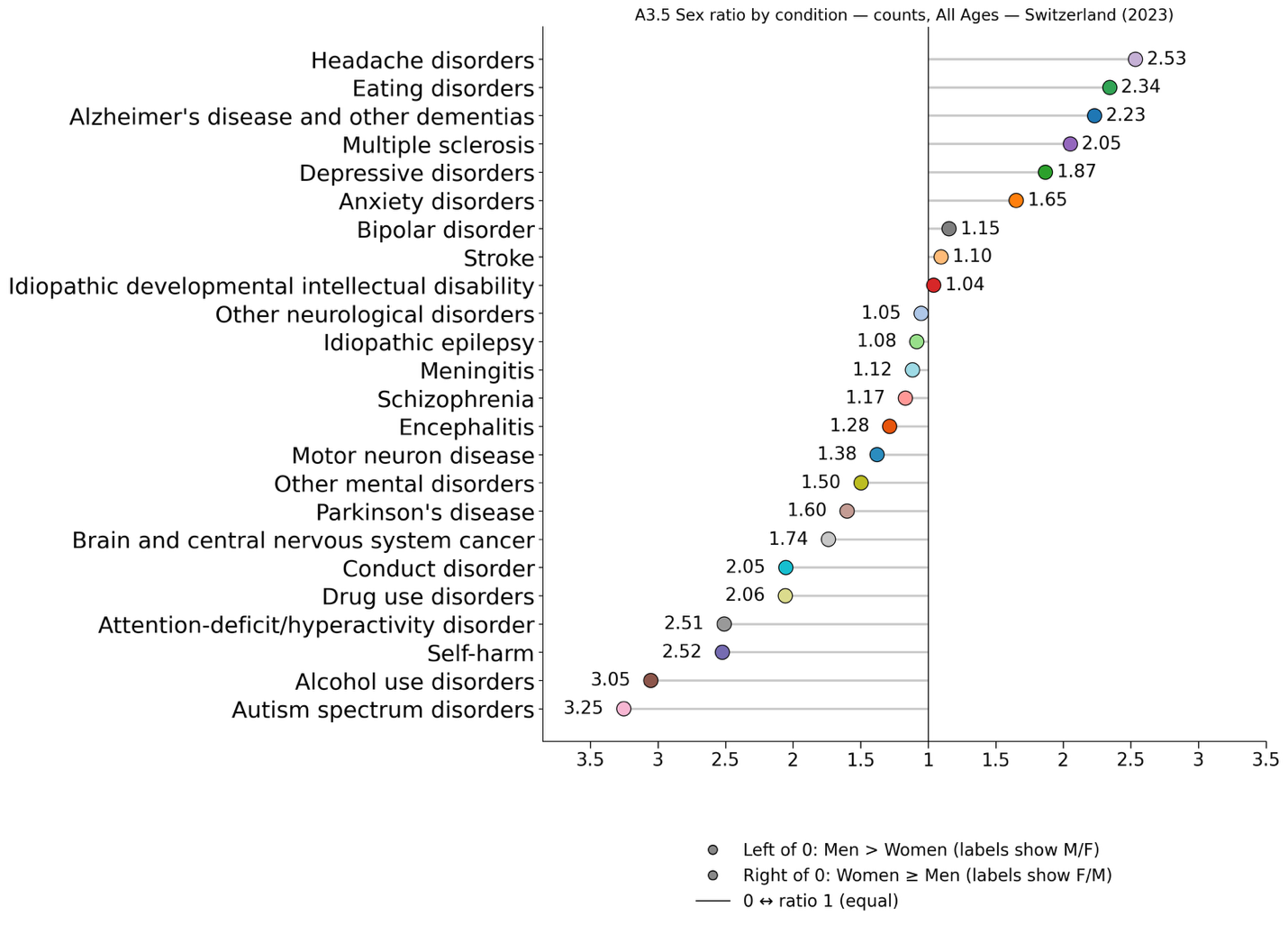
**Figure S2: Sex ratio of DALYs by brain disorder, Switzerland 2023**

Symmetric sex-ratio plot centered at 1 (equal burden). Points to the right indicate higher burden in women (labels show F/M), and points to the left indicate higher burden in men (labels show M/F). Female-to-male differences were largest for headache disorders (2.53 times higher in females), eating disorders (2.34 times higher) and Alzheimer’s disease and other dementias (2.23 times higher). By contrast, men experienced greater burden for autism spectrum disorders (3.25 times higher in males), alcohol use disorders (3.05 times higher) and self-harm (2.52 times higher). One dot per disorder; both sexes, all ages; DALYs=disability-adjusted life-years.

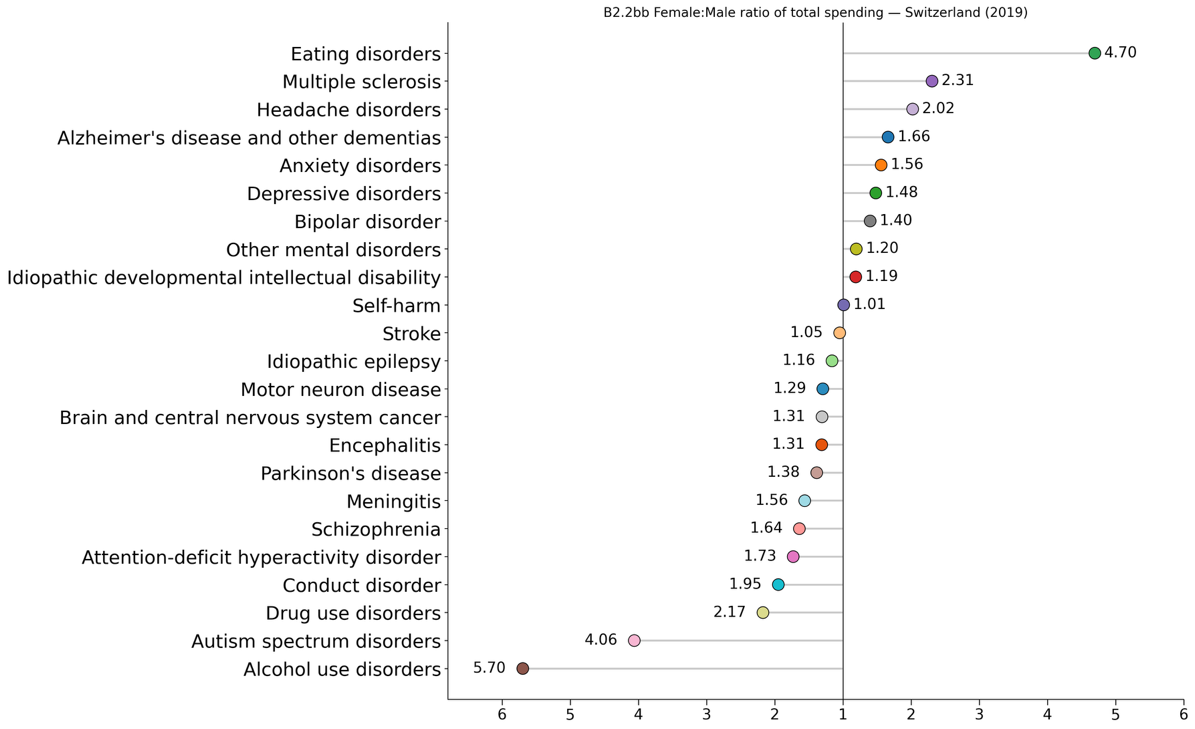

**Figure S3: Sex ratio of total direct health-care spending by brain disorder, Switzerland 2019**

Symmetric sex-ratio plot centered at 1 (equal spending). Points to the right indicate higher spending in women (labels show F/M), and points to the left indicate higher spending in men (labels show M/F). The largest female-to-male differences were observed for eating disorders (4.70 times higher in females), multiple sclerosis (2.31 times higher) and headache disorders (2.02 times higher). By contrast, men had higher total spending for alcohol use disorders (5.70 times higher in males), autism spectrum disorders (4.06 times higher), and drug use disorders (2.17 times higher). One dot per disorder; total spending across care types; all ages.

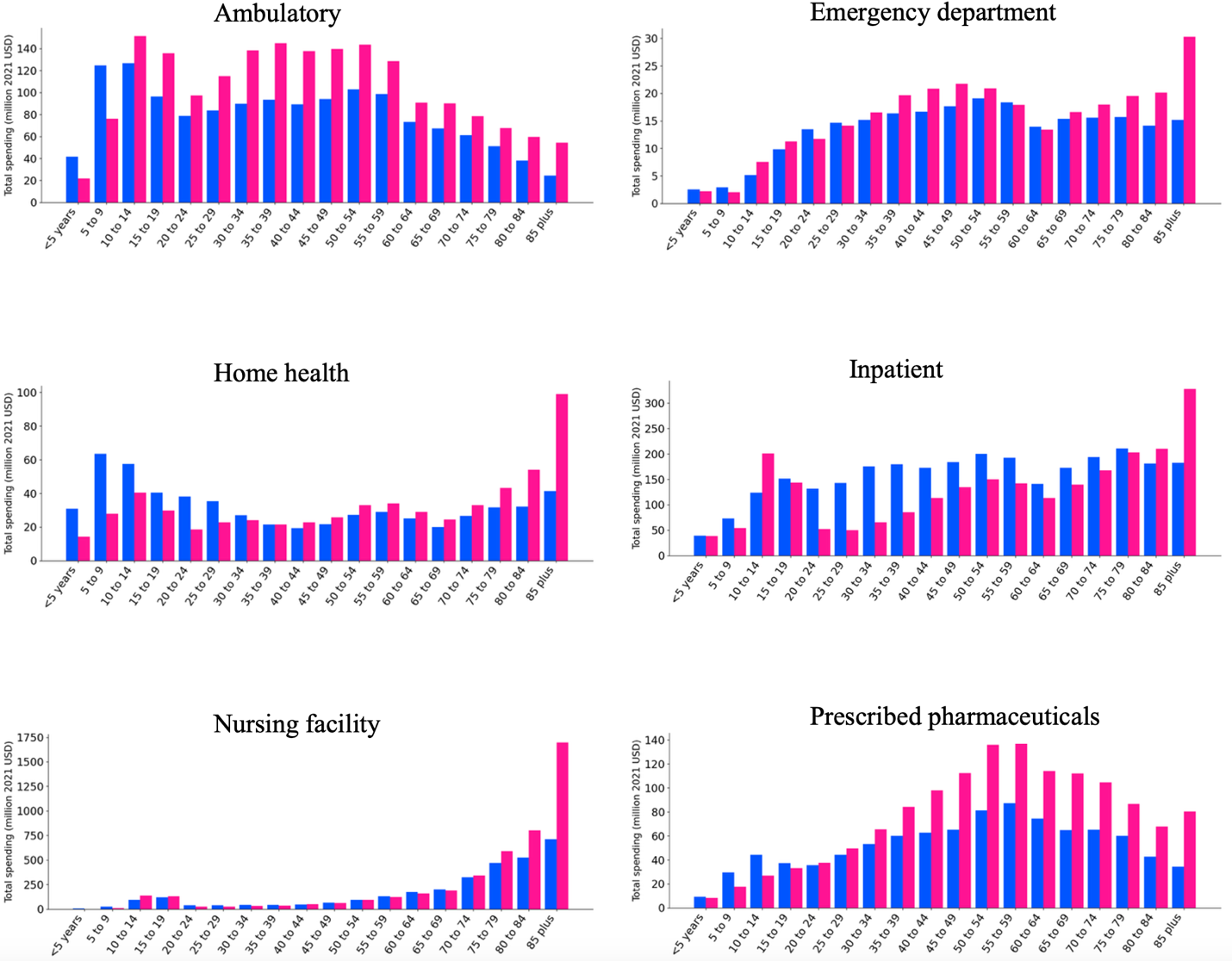

**Figure S4: Type of spending by age and sex, Switzerland 2019**

Total direct spending (million 2021 US$) on ambulatory, emergency department, home health, inpatient, nursing facility, prescribed pharmaceuticals, by 5-year age group; side-by-side bars for males (blue) and females (pink). All brain disorders combined.

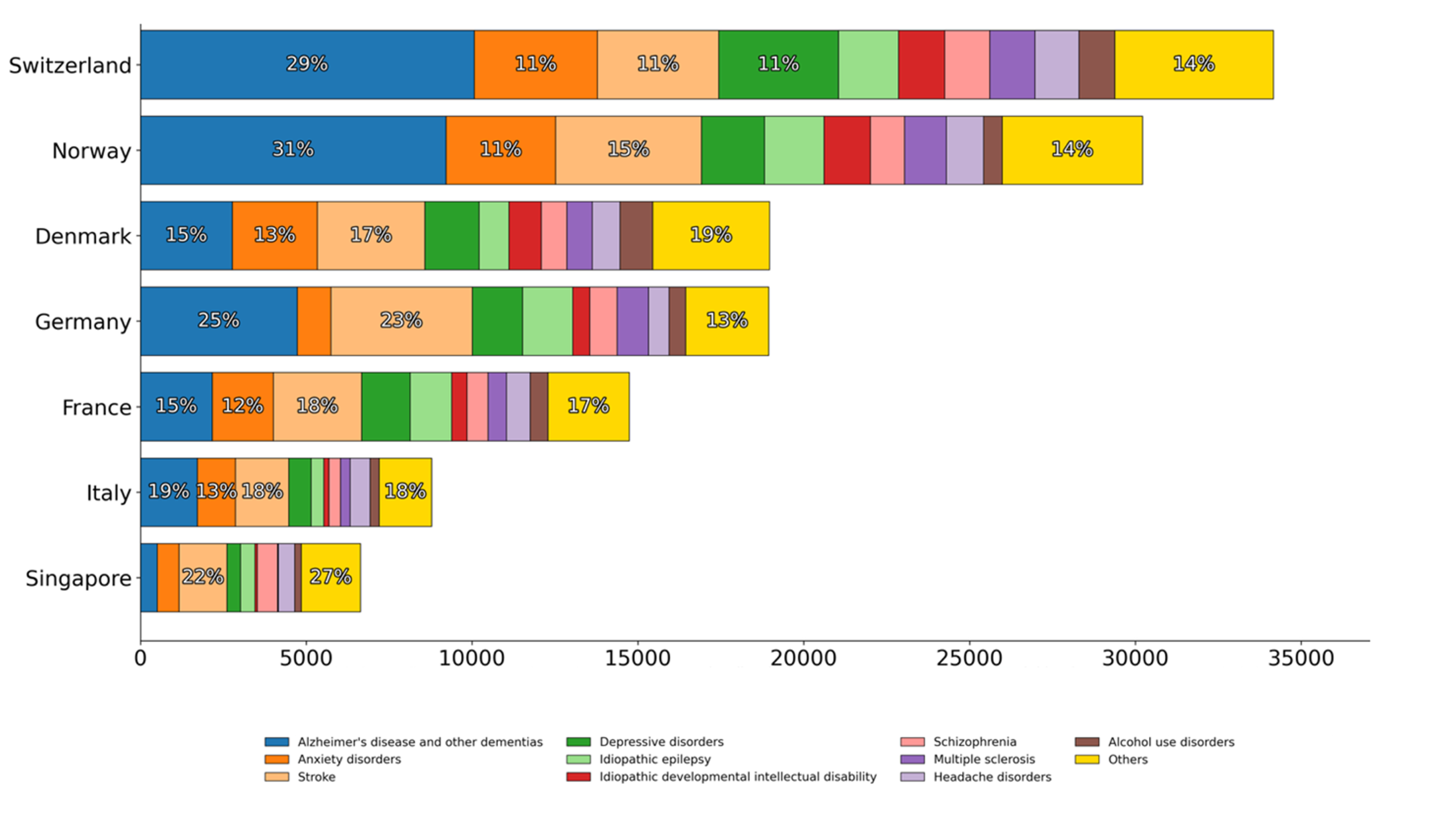

**Figure S5. Per-capita spending per DALY by condition and country, 2019**

Stacked horizontal bars for Switzerland, Norway, Germany, Denmark, France, Italy and Singapore. Each bar shows spending per DALY (2021 US$ per unit of burden) computed as per-capita spending (all ages)/ DALY rate (all ages) x 100 000. Segments denote condition contributions and sum to each country’s total; totals displayed at bar ends.

**Table S1: Health burden mean and uncertainty intervals for 24 brain disorders, Switzerland 2023**

| CONDITION | MEAN VALUE | LOWER | UPPER |
| --- | --- | --- | --- |
| Alzheimer’s disease and other dementias | 100691.4 | 44301.1 | 212704.0 |
| Depressive disorders | 94630.4 | 62904.5 | 135492.1 |
| Anxiety disorders | 84878.9 | 47268.4 | 146835.5 |
| Stroke | 70360.9 | 58629.1 | 79196.3 |
| Headache disorders | 58602.2 | 38364.3 | 83653.7 |
| Self-harm | 38772.0 | 34201.1 | 43436.3 |
| Alcohol use disorders | 29280.8 | 20242.8 | 41304.1 |
| Drug use disorders | 27784.8 | 21908.9 | 34523.4 |
| Schizophrenia | 20399.6 | 13477.9 | 28971.0 |
| Parkinson’s disease | 18198.5 | 15454.2 | 20369.6 |
| Brain and central nervous system cancer | 15976.1 | 13892.7 | 18087.7 |
| Autism spectrum disorders | 14153.4 | 6048.9 | 29936.4 |
| Other mental disorders | 13928.0 | 8805.7 | 20191.9 |
| Bipolar disorder | 13614.8 | 7564.3 | 21335.5 |
| Other neurological disorders | 11229.4 | 9091.0 | 13810.8 |
| Idiopathic epilepsy | 10327.2 | 5762.4 | 19637.9 |
| Eating disorders | 8257.1 | 4716.4 | 13306.6 |
| Multiple sclerosis | 7572.0 | 5897.1 | 9559.9 |
| Motor neuron disease | 6117.4 | 5242.1 | 6989.9 |
| Conduct disorder | 3703.4 | 2028.8 | 6214.5 |
| Attention-deficit/hyperactivity disorder | 3269.2 | 1748.0 | 5296.4 |
| Idiopathic developmental intellectual disability | 1939.7 | 612.5 | 4189.9 |
| Meningitis | 1176.3 | 985.6 | 1375.2 |
| Encephalitis | 1084.6 | 854.0 | 1368.5 |

*Data are mean disability-adjusted life-years (DALYs) with 95% uncertainty intervals, from the Global Burden of Disease 2023 for Switzerland.

**Table S2: Economic burden mean and uncertainty intervals for 23 brain disorders, Switzerland 2019**

| Condition | Value (million USD) | Lower (million USD) | Upper (million USD) |
| --- | --- | --- | --- |
| Alzheimer’s disease and other dementias | 5969.8 | 2540.9 | 15193.2 |
| Anxiety disorders | 2204.9 | 1910.4 | 2599.4 |
| Stroke | 2169.1 | 1428.4 | 3351.8 |
| Depressive disorders | 2141.7 | 1791.9 | 2634.8 |
| Idiopathic epilepsy | 1076.4 | 814.3 | 1476.6 |
| Idiopathic developmental intellectual disability | 822.2 | 533.9 | 1287.6 |
| Schizophrenia | 808.8 | 637.9 | 1117.5 |
| Multiple sclerosis | 806.2 | 538.8 | 1199.9 |
| Headache disorders | 789.1 | 632.2 | 1009.7 |
| Alcohol use disorders | 639.5 | 559.4 | 734.0 |
| Parkinson’s disease | 598.2 | 443.8 | 879.8 |
| Attention-deficit hyperactivity disorder | 343.7 | 262.2 | 435.1 |
| Autism spectrum disorders | 336.3 | 272.2 | 454.0 |
| Bipolar disorder | 329.7 | 245.5 | 439.6 |
| Brain and central nervous system cancer | 308.6 | 244.0 | 394.1 |
| Other mental disorders | 269.5 | 247.0 | 292.5 |
| Drug use disorders | 221.7 | 198.7 | 245.1 |
| Conduct disorder | 108.6 | 94.5 | 128.1 |
| Meningitis | 86.4 | 62.4 | 129.2 |
| Motor neuron disease | 71.8 | 52.5 | 109.4 |
| Encephalitis | 58.5 | 40.2 | 91.0 |
| Eating disorders | 56.4 | 50.3 | 62.6 |
| Self-harm | 48.6 | 42.1 | 56.9 |

*Data are total direct health-care spending (mean and 95% uncertainty intervals) for Switzerland in 2019, expressed in millions of 2021 USD. Estimates are from the Institute for Health Metrics and Evaluation (IHME) direct health-spending database.

**Table S3. Reverse-regression results (dependent variable: log DALYs)**

|  | **Model 1** | **Model 2** |
| --- | --- | --- |
| Time window | 1 year | 5 years |
| log(Spending) | 0.025 (0.018) | 0.067 (0.059) |
| log(DALY) | 0.91 (0.021)** | 0.76 (0.052)** |
| Observations (conditions x years) | 437 (23 x 19) | 345 (23 x 15) |
| R² | 1.000 | 1.000 |
| Adjusted R² | 1.000 | 1.000 |
| Abbreviations: DALY, disability-adjusted life-years. | | |
| Notes: Standard errors clustered by condition. Coefficients should be interpreted as elasticities. Constants are excluded. * p<0.05; ** p<0.001. | | |

**Table S4. Coefficients for disorder and year fixed effects in the one-year lag spending model**

|  | **Beta** | **SE** | **p** |
| --- | --- | --- | --- |
| **Condition fixed effects** | | | |
| Alzheimer’s disease and other dementias | 0.1073 | 0.051 | 0.036* |
| Anxiety disorders | 0.0570 | 0.027 | 0.036* |
| Attention-deficit hyperactivity disorder | 0.1365 | 0.135 | 0.312 |
| Autism spectrum disorders | 0.0241 | 0.040 | 0.552 |
| Bipolar disorder | 0.0315 | 0.047 | 0.501 |
| Brain and central nervous system cancer | 0.0101 | 0.033 | 0.760 |
| Conduct disorder | 0.0448 | 0.103 | 0.665 |
| Depressive disorders | 0.0355 | 0.031 | 0.257 |
| Drug use disorders | -0.0781 | 0.014 | 0.000* |
| Eating disorders | -0.0655 | 0.060 | 0.272 |
| Encephalitis | 0.1211 | 0.166 | 0.465 |
| Headache disorders | -0.0203 | 0.023 | 0.385 |
| Idiopathic developmental intellectual disability | 0.2468 | 0.182 | 0.174 |
| Idiopathic epilepsy | 0.1385 | 0.093 | 0.137 |
| Meningitis | 0.0702 | 0.125 | 0.574 |
| Motor neuron disease | -0.0250 | 0.074 | 0.735 |
| Multiple sclerosis | 0.1437 | 0.104 | 0.168 |
| Other mental disorders | 0.0179 | 0.045 | 0.693 |
| Parkinson’s disease | 0.0818 | 0.057 | 0.154 |
| Schizophrenia | 0.0609 | 0.041 | 0.136 |
| Self-harm | -0.2343 | 0.089 | 0.008* |
| Stroke | 0.0250 | 0.037 | 0.499 |
| **Year fixed effects** |  |  |  |
| 2002 | 0.0400 | 0.007 | 0.000* |
| 2003 | 0.0248 | 0.007 | 0.001* |
| 2004 | 0.0104 | 0.007 | 0.147 |
| 2005 | -0.0105 | 0.006 | 0.065 |
| 2006 | -0.0417 | 0.010 | 0.000* |
| 2007 | 0.0010 | 0.011 | 0.931 |
| 2008 | 0.0054 | 0.008 | 0.522 |
| 2009 | 0.0085 | 0.008 | 0.272 |
| 2010 | -0.0218 | 0.011 | 0.051 |
| 2011 | 0.0041 | 0.010 | 0.676 |
| 2012 | 0.0055 | 0.017 | 0.750 |
| 2013 | 0.0159 | 0.012 | 0.186 |
| 2014 | 0.0201 | 0.012 | 0.102 |
| 2015 | 0.0389 | 0.010 | 0.000* |
| 2016 | 0.0487 | 0.011 | 0.000* |
| 2017 | 0.0600 | 0.012 | 0.000* |
| 2018 | 0.0536 | 0.013 | 0.000* |
| 2019 | 0.0458 | 0.014 | 0.001* |

**Table S5. Coefficients for disorder and year fixed effects in the five-year lag spending model**

|  | **Beta** | **SE** | **p** |
| --- | --- | --- | --- |
| **Condition fixed effects** | | | |
| Alzheimer’s disease and other dementias | 0.1535 | 0.226 | 0.497 |
| Anxiety disorders | 0.0951 | 0.121 | 0.430 |
| Attention-deficit hyperactivity disorder | 0.3011 | 0.421 | 0.474 |
| Autism spectrum disorders | 0.1075 | 0.142 | 0.448 |
| Bipolar disorder | 0.0878 | 0.151 | 0.559 |
| Brain and central nervous system cancer | 0.0386 | 0.110 | 0.726 |
| Conduct disorder | 0.1354 | 0.339 | 0.689 |
| Depressive disorders | 0.0187 | 0.136 | 0.891 |
| Drug use disorders | -0.1785 | 0.059 | 0.002* |
| Eating disorders | -0.0352 | 0.254 | 0.890 |
| Encephalitis | 0.4129 | 0.554 | 0.456 |
| Headache disorders | -0.0834 | 0.083 | 0.315 |
| Idiopathic developmental intellectual disability | 0.5359 | 0.562 | 0.341 |
| Idiopathic epilepsy | 0.2902 | 0.289 | 0.316 |
| Meningitis | 0.1846 | 0.412 | 0.654 |
| Motor neuron disease | 0.0195 | 0.269 | 0.942 |
| Multiple sclerosis | 0.3265 | 0.323 | 0.312 |
| Other mental disorders | 0.0528 | 0.147 | 0.720 |
| Parkinson’s disease | 0.1953 | 0.180 | 0.279 |
| Schizophrenia | 0.1222 | 0.127 | 0.338 |
| Self-harm | -0.4713 | 0.296 | 0.112 |
| Stroke | -0.0116 | 0.155 | 0.941 |
| **Year fixed effects** |  |  |  |
| 2006 | -0.0551 | 0.010 | 0.000* |
| 2007 | -0.0563 | 0.014 | 0.000* |
| 2008 | -0.0460 | 0.010 | 0.000* |
| 2009 | -0.0295 | 0.013 | 0.027* |
| 2010 | -0.0444 | 0.013 | 0.001* |
| 2011 | -0.0305 | 0.016 | 0.050 |
| 2012 | -0.0242 | 0.022 | 0.276 |
| 2013 | -0.0087 | 0.028 | 0.759 |
| 2014 | 0.0081 | 0.032 | 0.800 |
| 2015 | 0.0403 | 0.030 | 0.178 |
| 2016 | 0.0698 | 0.029 | 0.015* |
| 2017 | 0.1003 | 0.023 | 0.000* |
| 2018 | 0.1128 | 0.024 | 0.000* |
| 2019 | 0.1097 | 0.029 | 0.000* |
